# Adherence to the PRISMA statement and risk of bias assessment in Systematic Reviews in rehabilitation journals: a protocol for a meta-research study

**DOI:** 10.1101/2021.01.21.21250260

**Authors:** Tiziano Innocenti, Daniel Feller, Silvia Giagio, Stefano Salvioli, Silvia Minnucci, Fabrizio Brindisino, Alessandro Chiarotto, Raymond Ostelo

**Author notes:** **Corresponding author:** Dr. Tiziano Innocenti, Department of Health Sciences, Faculty of Science, VU University, Amsterdam Movement Sciences, The Netherlands.

## Abstract

**Objective:** The aim of this study will be to assess the adherence to the reporting quality standards set forth in the PRISMA Statement checklist of a random sample of systematic reviews (SRs) published in rehabilitation journals, and to assess the association between this adherence and the risk of bias of these SRs.

**Methods and Analysis:** A cross-sectional analysis is planned on a random sample of 200 SRs published between 2011and 2020 in the 68 journals indexed under “rehabilitation” category in InCites Journal Citation Report.

Randomization will be stratified by publication date and journal ranking (quartile range; Q1-2 and Q3-4) to include an equal number of studies from 2011 to 2015 (Q1-Q2=50 and Q3-Q4=50) and from 2016 to 2020 (Q1-Q2=50 and Q3-Q4=50). SRs (with or without meta-analysis) published between 2011 and 2020 as full-text scientific articles in the 68 rehabilitation journals will be included. Narrative reviews, mixed-methods reviews, meta-ethnography reviews, umbrella reviews, scoping reviews, editorials, letters and news reports will be excluded. The primary analysis will address the completeness of the reporting for each study and the relationship between PRISMA adherence and risk of bias. This will be a descriptive analysis through descriptive statistics and graphical representation.

**Ethics and Dissemination:** Several studies have shown the positive influence of reporting guidelines on the completeness of research reporting but no one investigated the use and the appropriateness of reporting guidelines in physical therapy research. Therefore, this study will add relevant knowledge that may contribute to improve further the reporting of rehabilitation research. The results of this research will be published in a peer-reviewed journal and will be presented at relevant (inter)national scientific events.

## Introduction

Research in health sciences should have the potential to advance scientific understanding, or to improve the treatment or prevention of disease. Clinical practice and public health policy decisions should be based on high-quality research findings^1^. The purpose of a research report (e.g. a scientific article) is to communicate the design, execution, and findings of a study with precision and accuracy^2^; this is relevant for several stakeholder groups, including researchers, clinicians, systematic reviewers and patients^3^. Research reports should include all relevant information about methods and results to be useful to all these categories of potential readers. Additionally, accurate reporting of a study is essential to judge its validity and the clinical applicability of its findings^1^.

Problems about reporting of a scientific study can affect research in different ways. For example, it is known that study methods are frequently not described in adequate detail and that results are presented ambiguously, incompletely or selectively^3^. The consequence is that many reports cannot be used for replication studies, or they are even harmful, as well as a waste of resources^4^. To overcome these problems, reporting guidelines (RGs) have been developed to support authors in reporting research methods and results.

In 2014, twenty-eight rehabilitation journals have simultaneously published an editorial to highlight the need of using RGs to ensure the quality and the completeness of rehabilitation research^5^. Chan and colleagues concluded the editorial hoping “…that simultaneous implementation of this new reporting requirement will send a strong message to all disability and rehabilitation researchers about the need to adhere to the highest standards when performing and disseminating research.” Systematic reviews (SRs) and meta-analyses provide the highest quality of scientific evidence, and are important resources in shaping our clinical decision-making^6^. They represent an efficient way for clinicians to keep up-to-date with the current state of evidence and provide a starting point for development of clinical guidelines^7, 8^. Therefore, adequate reporting of the methodology and the results is essential for evaluating the strengths and weaknesses of the evidence provided^9^. Moreover, complete, and transparent reported research aids reproducibility and critical appraisal^10^. Indeed, critical appraisal of study methods enables judgements of the degree to which results and author interpretations overestimate or under-estimate study effects, and is highly dependent on what is reported within research reports. For example, if a study has better reporting, this could positively impact the evaluation of risk of bias^11^.

The lack of standardization and poor quality of reporting among systematic reviews and meta-analyses had led to the development of the QUOROM statement (Quality of Reporting of Meta-analyses) in 1999^12^, and its subsequent evolution into the PRISMA statement (Preferred Reporting Items for Systematic Reviews and Meta-analyses) in 2009^9^, that is an evidence-based minimum set of items for reporting SRs and meta-analyses. This statement focuses on the reporting of reviews that evaluate randomised controlled trials but can also be used for reporting SRs of other types of research. It consists of 27 items that pertain to the contents of a SR and meta-analysis, including the title, abstract, methods, results, discussion and funding. PRISMA statement was update in 2020, but only a preprint version is available^13^.

Regarding the risk of bias in SRs, a tool (ROBIS Tool) specifically designed to assess the risk of bias in systematic reviews was launched in 2016^14^. ROBIS follows a domain-based approach and covers not only evaluation of the internal validity of the review process but also the relevance of the review question for its users. It comprises four specific domains (eligibility criteria, identification and selection of studies, data collection and study appraisal, synthesis and findings) and an overall risk of bias judgment. Each domain has several signaling questions to help the reader formulate judgments. The judgment about risk of bias can be ‘‘high,’’ ‘‘low,’’ or ‘‘unclear’’.

Many previous studies assessed the adherence to PRISMA checklist in SRs in the medical field^10, 15^; in the field of orthopaedic surgery, one study^16^ analysed the methodological quality of relevant studies which were published in the top five highest impact factor orthopaedic journals and found that only 68% of items within the PRISMA statement were reported. Little is known in rehabilitation field: a previus study^17^ confirmed that most of the SRs authors (∼ 65%) publishing in high impact rehabilitation journals did not mention their use and approximately 40% of those who declared using RGs did not do it in an appropriate manner. This occurred despite many rehabilitation journals requiring RGs as mandatory during submission.

To our knowledge, the adherence to the PRISMA checklist and the relationship between risk of bias and the completeness of the reporting in systematic review of rehabilitation intervention has not been systematically evaluated.

The aim of this study will be to assess the adherence to the reporting quality standards set forth in the PRISMA Statement checklist of a random sample of SRs published in rehabilitation journals, and to assess the association between this adherence and the risk of bias of these SRs.

### Objectives

#### Primary Objectives

1. To evaluate the completeness of the reporting in SRs published in rehabilitation journals between 2011 and 2020 by adherence to indications contained in PRISMA checklist^9^.
2. To investigate the possible relationship between completeness of reporting and risk of bias.

#### Secondary Objectives

1. To compare the adherence to the PRISMA checklist^9^ and year of publication, journal proprieties (e.g. Quartile, Open Access/Subscription) and others factors (e.g. registration (yes/no) of the study protocol) in order to investigate the impact of these features.

## METHODS AND ANALYSIS

We will conduct a cross-sectional analysis in a random sample of 200 SRs published between 2011 and 2020 in the 68 journals indexed under “rehabilitation” category in InCites Journal Citation Report^18^. This sampling was chosen to include all the journals in the rehabilitation field. The 68 rehabilitation journals indexed in InCites are reported in Table 1.

**Table 1:**
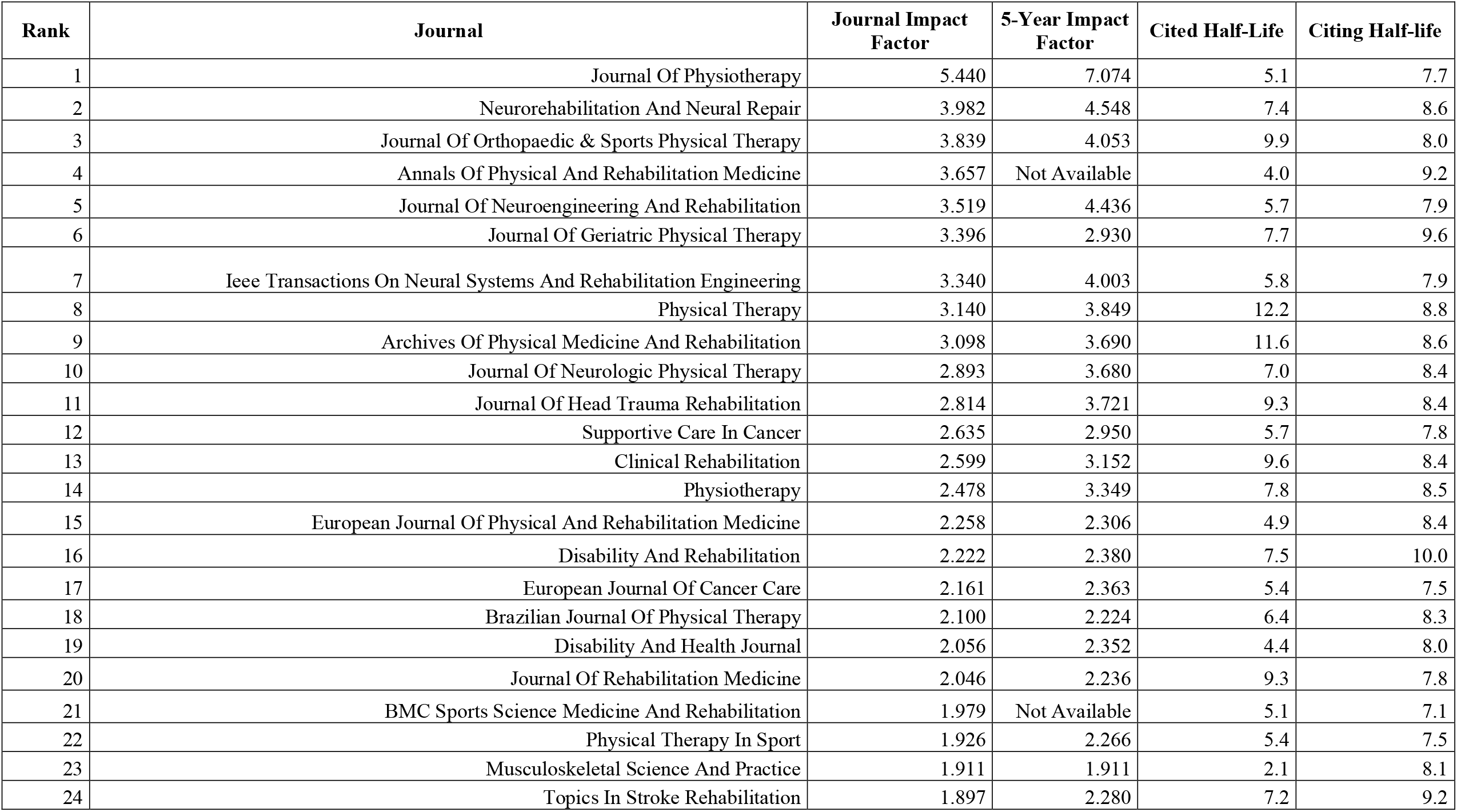
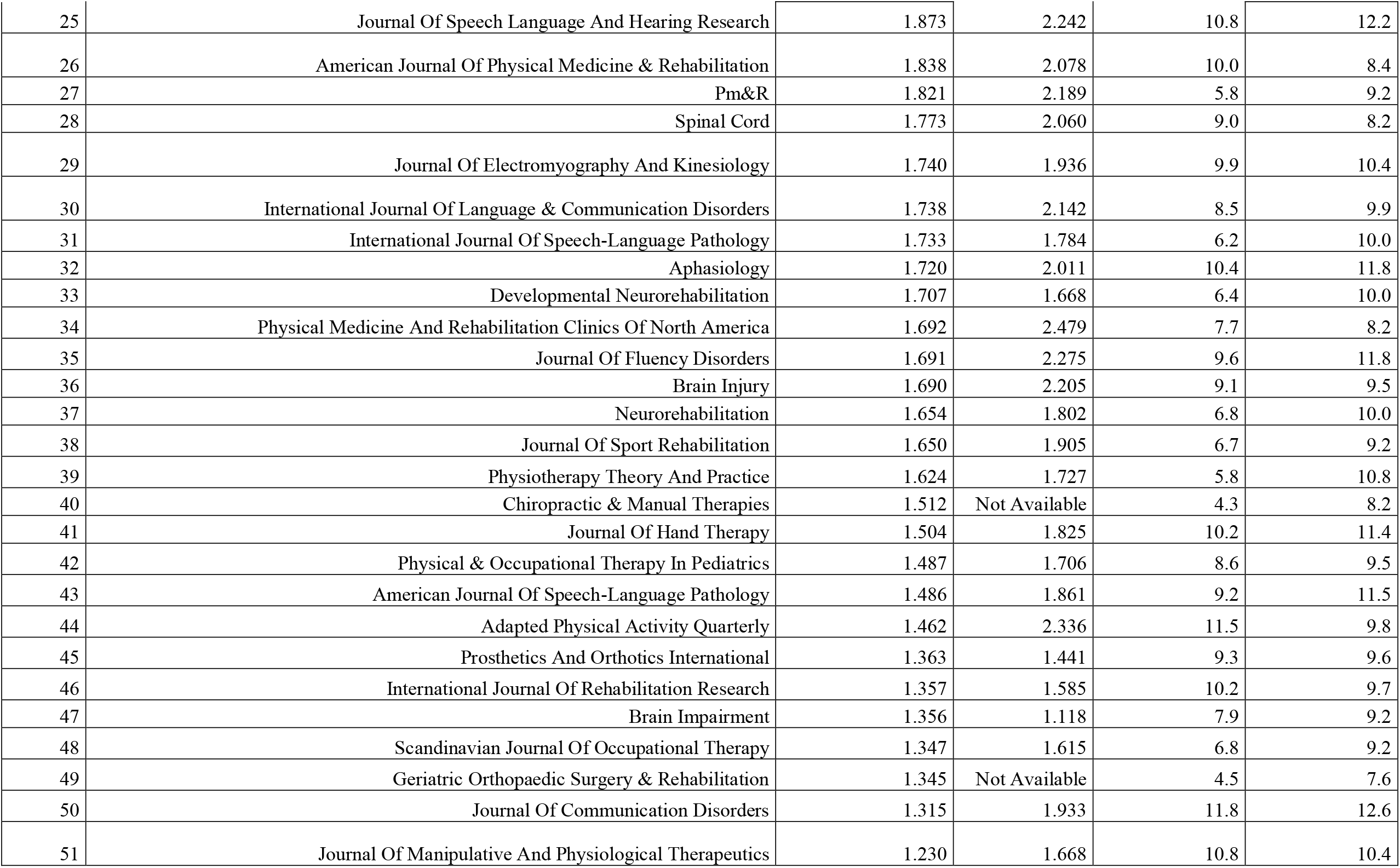
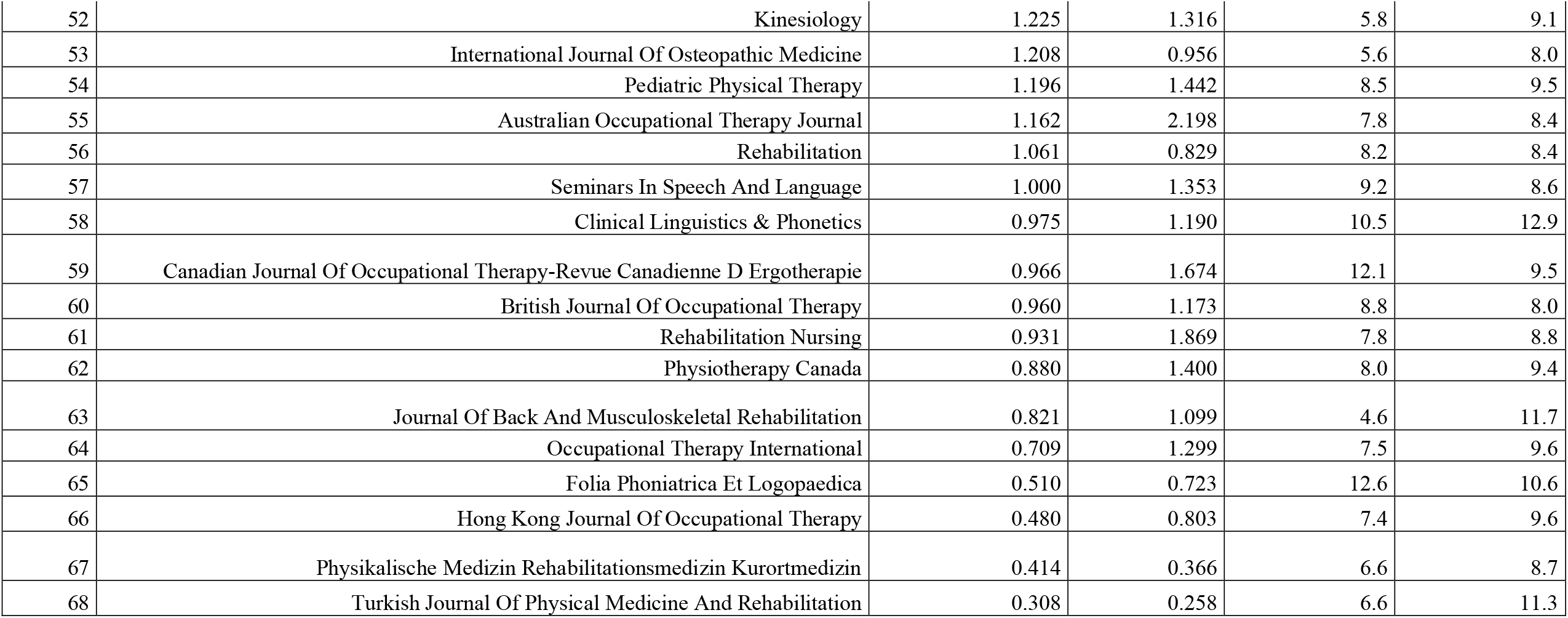
Journals selected (Year 2019 according InCites Journal Citation Reports)

| Rank | Journal | Journal Impact Factor | 5-Year Impact Factor | Cited Half-Life | Citing Half-life |
| --- | --- | --- | --- | --- | --- |
| 1 | Journal Of Physiotherapy | 5.440 | 7.074 | 5.1 | 7.7 |
| 2 | Neurorehabilitation And Neural Repair | 3.982 | 4.548 | 7.4 | 8.6 |
| 3 | Journal Of Orthopaedic & Sports Physical Therapy | 3.839 | 4.053 | 9.9 | 8.0 |
| 4 | Annals Of Physical And Rehabilitation Medicine | 3.657 | Not Available | 4.0 | 9.2 |
| 5 | Journal Of Neuroengineering And Rehabilitation | 3.519 | 4.436 | 5.7 | 7.9 |
| 6 | Journal Of Geriatric Physical Therapy | 3.396 | 2.930 | 7.7 | 9.6 |
| 7 | Ieee Transactions On Neural Systems And Rehabilitation Engineering | 3.340 | 4.003 | 5.8 | 7.9 |
| 8 | Physical Therapy | 3.140 | 3.849 | 12.2 | 8.8 |
| 9 | Archives Of Physical Medicine And Rehabilitation | 3.098 | 3.690 | 11.6 | 8.6 |
| 10 | Journal Of Neurologic Physical Therapy | 2.893 | 3.680 | 7.0 | 8.4 |
| 11 | Journal Of Head Trauma Rehabilitation | 2.814 | 3.721 | 9.3 | 8.4 |
| 12 | Supportive Care In Cancer | 2.635 | 2.950 | 5.7 | 7.8 |
| 13 | Clinical Rehabilitation | 2.599 | 3.152 | 9.6 | 8.4 |
| 14 | Physiotherapy | 2.478 | 3.349 | 7.8 | 8.5 |
| 15 | European Journal Of Physical And Rehabilitation Medicine | 2.258 | 2.306 | 4.9 | 8.4 |
| 16 | Disability And Rehabilitation | 2.222 | 2.380 | 7.5 | 10.0 |
| 17 | European Journal Of Cancer Care | 2.161 | 2.363 | 5.4 | 7.5 |
| 18 | Brazilian Journal Of Physical Therapy | 2.100 | 2.224 | 6.4 | 8.3 |
| 19 | Disability And Health Journal | 2.056 | 2.352 | 4.4 | 8.0 |
| 20 | Journal Of Rehabilitation Medicine | 2.046 | 2.236 | 9.3 | 7.8 |
| 21 | BMC Sports Science Medicine And Rehabilitation | 1.979 | Not Available | 5.1 | 7.1 |
| 22 | Physical Therapy In Sport | 1.926 | 2.266 | 5.4 | 7.5 |
| 23 | Musculoskeletal Science And Practice | 1.911 | 1.911 | 2.1 | 8.1 |
| 24 | Topics In Stroke Rehabilitation | 1.897 | 2.280 | 7.2 | 9.2 |
| 25 | Journal Of Speech Language And Hearing Research | 1.873 | 2.242 | 10.8 | 12.2 |
| 26 | American Journal Of Physical Medicine & Rehabilitation | 1.838 | 2.078 | 10.0 | 8.4 |
| 27 | Pm&R | 1.821 | 2.189 | 5.8 | 9.2 |
| 28 | Spinal Cord | 1.773 | 2.060 | 9.0 | 8.2 |
| 29 | Journal Of Electromyography And Kinesiology | 1.740 | 1.936 | 9.9 | 10.4 |
| 30 | International Journal Of Language & Communication Disorders | 1.738 | 2.142 | 8.5 | 9.9 |
| 31 | International Journal Of Speech-Language Pathology | 1.733 | 1.784 | 6.2 | 10.0 |
| 32 | Aphasiology | 1.720 | 2.011 | 10.4 | 11.8 |
| 33 | Developmental Neurorehabilitation | 1.707 | 1.668 | 6.4 | 10.0 |
| 34 | Physical Medicine And Rehabilitation Clinics Of North America | 1.692 | 2.479 | 7.7 | 8.2 |
| 35 | Journal Of Fluency Disorders | 1.691 | 2.275 | 9.6 | 11.8 |
| 36 | Brain Injury | 1.690 | 2.205 | 9.1 | 9.5 |
| 37 | Neurorehabilitation | 1.654 | 1.802 | 6.8 | 10.0 |
| 38 | Journal Of Sport Rehabilitation | 1.650 | 1.905 | 6.7 | 9.2 |
| 39 | Physiotherapy Theory And Practice | 1.624 | 1.727 | 5.8 | 10.8 |
| 40 | Chiropractic & Manual Therapies | 1.512 | Not Available | 4.3 | 8.2 |
| 41 | Journal Of Hand Therapy | 1.504 | 1.825 | 10.2 | 11.4 |
| 42 | Physical & Occupational Therapy In Pediatrics | 1.487 | 1.706 | 8.6 | 9.5 |
| 43 | American Journal Of Speech-Language Pathology | 1.486 | 1.861 | 9.2 | 11.5 |
| 44 | Adapted Physical Activity Quarterly | 1.462 | 2.336 | 11.5 | 9.8 |
| 45 | Prosthetics And Orthotics International | 1.363 | 1.441 | 9.3 | 9.6 |
| 46 | International Journal Of Rehabilitation Research | 1.357 | 1.585 | 10.2 | 9.7 |
| 47 | Brain Impairment | 1.356 | 1.118 | 7.9 | 9.2 |
| 48 | Scandinavian Journal Of Occupational Therapy | 1.347 | 1.615 | 6.8 | 9.2 |
| 49 | Geriatric Orthopaedic Surgery & Rehabilitation | 1.345 | Not Available | 4.5 | 7.6 |
| 50 | Journal Of Communication Disorders | 1.315 | 1.933 | 11.8 | 12.6 |
| 51 | Journal Of Manipulative And Physiological Therapeutics | 1.230 | 1.668 | 10.8 | 10.4 |
| 52 | Kinesiology | 1.225 | 1.316 | 5.8 | 9.1 |
| 53 | International Journal Of Osteopathic Medicine | 1.208 | 0.956 | 5.6 | 8.0 |
| 54 | Pediatric Physical Therapy | 1.196 | 1.442 | 8.5 | 9.5 |
| 55 | Australian Occupational Therapy Journal | 1.162 | 2.198 | 7.8 | 8.4 |
| 56 | Rehabilitation | 1.061 | 0.829 | 8.2 | 8.4 |
| 57 | Seminars In Speech And Language | 1.000 | 1.353 | 9.2 | 8.6 |
| 58 | Clinical Linguistics & Phonetics | 0.975 | 1.190 | 10.5 | 12.9 |
| 59 | Canadian Journal Of Occupational Therapy-Revue Canadienne D Ergotherapie | 0.966 | 1.674 | 12.1 | 9.5 |
| 60 | British Journal Of Occupational Therapy | 0.960 | 1.173 | 8.8 | 8.0 |
| 61 | Rehabilitation Nursing | 0.931 | 1.869 | 7.8 | 8.8 |
| 62 | Physiotherapy Canada | 0.880 | 1.400 | 8.0 | 9.4 |
| 63 | Journal Of Back And Musculoskeletal Rehabilitation | 0.821 | 1.099 | 4.6 | 11.7 |
| 64 | Occupational Therapy International | 0.709 | 1.299 | 7.5 | 9.6 |
| 65 | Folia Phoniatica Et Logopaedica | 0.510 | 0.723 | 12.6 | 10.6 |
| 66 | Hong Kong Journal Of Occupational Therapy | 0.480 | 0.803 | 7.4 | 9.6 |
| 67 | Physikalische Medizin Rehabilitationsmedizin Kurortmedizin | 0.414 | 0.366 | 6.6 | 8.7 |
| 68 | Turkish Journal Of Physical Medicine And Rehabilitation | 0.308 | 0.258 | 6.6 | 11.3 |

### Study selection criteria

SRs (with or without meta-analysis) published between 2011 and 2020 as full-text scientific articles in the 68 rehabilitation journals will be included. A standard or consensus definition of a systematic review does not exist^19^, so we will consider a review as systematic if include the following^20^:

- A defined research question
- Sources that were searched
- Inclusion and exclusion criteria and selection methods
- Critically appraises and reports the quality/risk of bias of the included studies
- Information about data analysis and synthesis

Narrative reviews, mixed-methods reviews, meta-ethnography reviews, umbrella reviews, scoping reviews, editorials, letters and news reports will be excluded.

### Study selection process

Journal tags for the journals will be identified in Medline and a detailed search strategy will be created to find all SRs published from 2011 to 2020 in this database (See Table 2). The searching will be performed only in Medline because all of the journals are indexed in Medline.

**Table 2:**
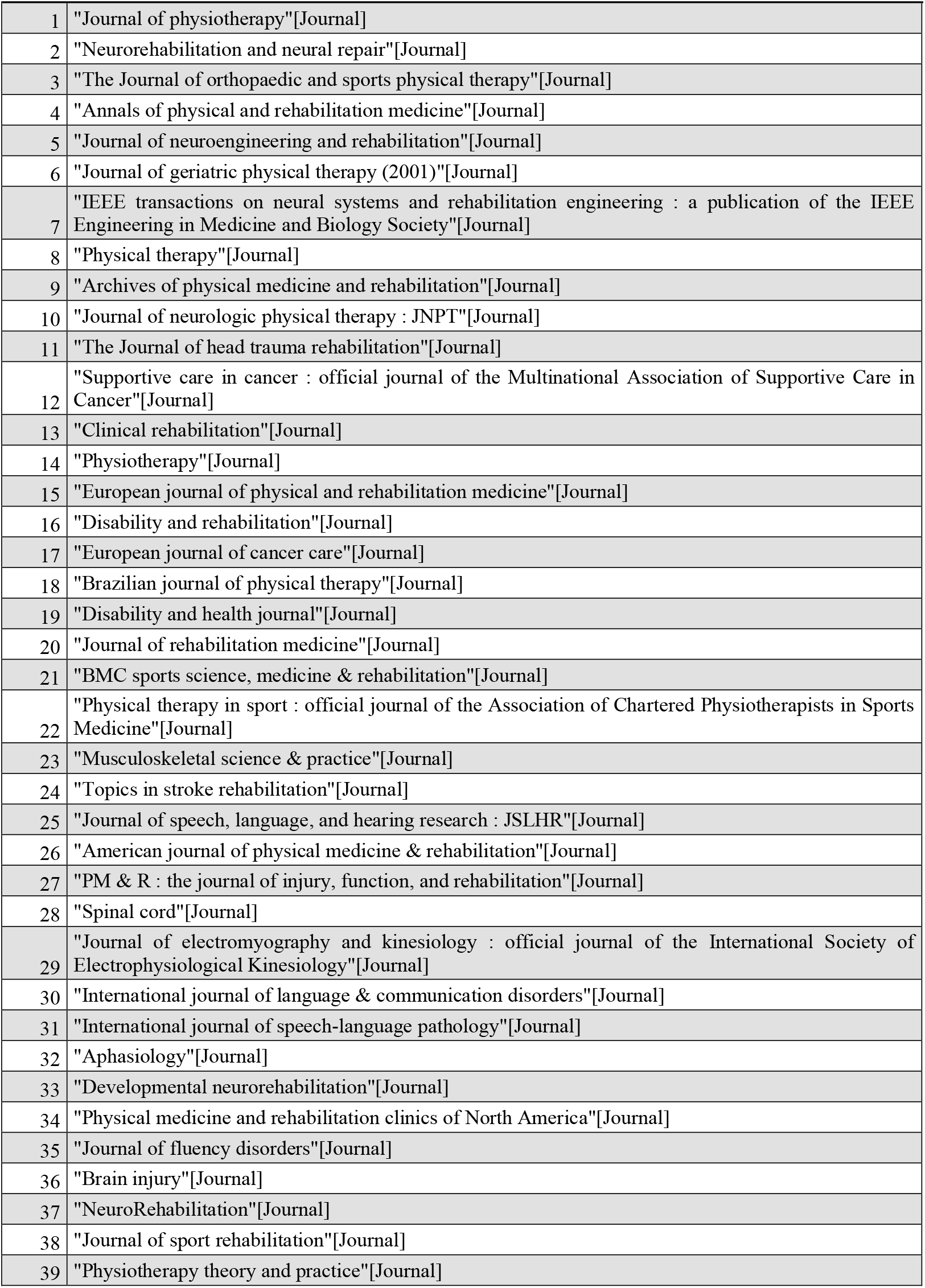
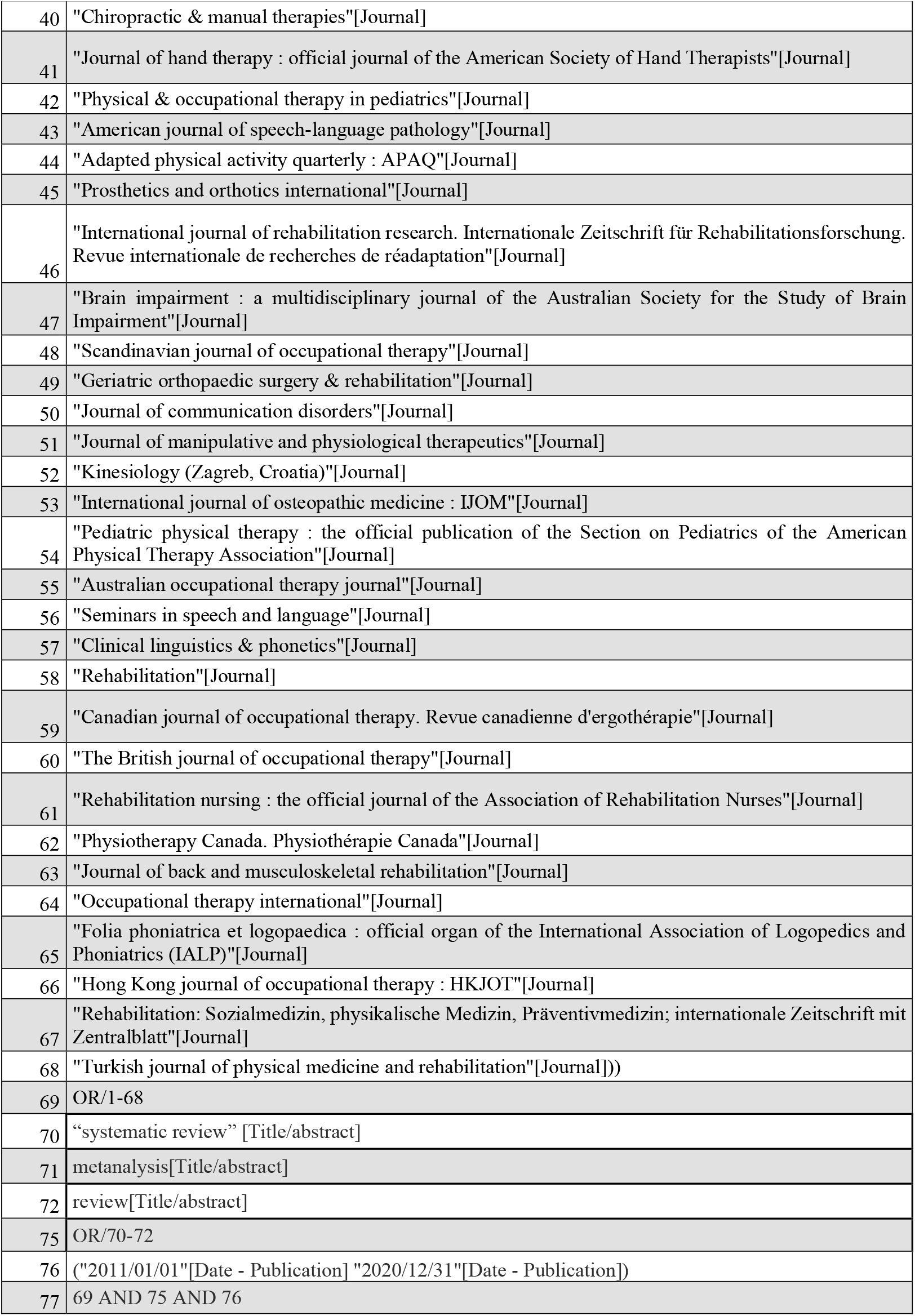
Search Strategy.

Titles and abstracts of the selected articles will be screened for eligibility in a double blinded process by two independent reviewers who select potentially eligible articles independently and based on selection criteria. Any disagreement will be resolved by a third reviewer. Subsequently, the study sample of 200 hits will be selected using a computerized random sequence generator^21^. Randomization will be stratified by publication date and journal ranking (quartile range; Q1-2 and Q3-4) to include an equal number of studies from 2011 to 2015 (Q1-Q2=50 and Q3-Q4=50) and from 2016 to 2020 ((Q1-Q2=50 and Q3-Q4=50). Study selection process will be summarized in a tailored flow diagram.

### Data extraction

Full-texts will be stored in EndNote X7 (Thomas Reuters, Philadelphia, Pennsylvania, USA). A data extraction form will be used by two reviewers and the following data will be extracted:

- First author and year of publication;
- Journal with characteristic (Quartile Range, Publication options - Open Access/subscription/Hybrid – mandatory use of RGs);
- Country in which was conducted;
- Rehabilitation research field (e.g. musculoskeletal, neurological, pediatrics, pelvic-floor disfunction, cardio-pulmonary, other)
- Published protocol (Yes/No)
- Adherence to the PRISMA checklist (see later);
- Risk of bias with the ROBIS checklist (see later).

Data extraction and the assessment of risk of bias from each SR will be performed by two reviewers, independently; when necessary, a third researcher will solve disagreements.

### Assessing the completeness of the reporting

We will determine compliance to each item of the PRISMA checklist^9^ through the use of 27 item checklist. Discrepancies between authors over application of checklist items will be resolved by consulting the published explanation of the PRISMA Statement^9^. Prior to scoring of study inter-rater agreement for each item on the 27-item checklist will be evaluated in a pilot study of 20 SRs and scored independently by three authors with post-graduate training in epidemiology and critical appraisal.

According to the explanation and elaboration statements of PRISMA checklist, each item will be marked with “1” if it was well described, incomplete or missing items with “0”, and not applicable items with “NA”. Total adherence for each item between studies and total adherence to the PRISMA Checklist for each study will be calculate.

Given the aim of this study will be to investigate the comprehensiveness of the reporting, authors of included studies will be not contacted for information omitted from manuscripts. Summary tables and graphics of extracted data of all included studies and a narrative synthesis will be provided.

### Assessing the risk of bias

ROBIS tool^14^ will be used to assess the risk of bias in the included studies. For each SR included a risk of bias judgment will be reached for each domain (1.concerns regarding specification of study; 2. concerns regarding methods used to identify and/or select studies eligibility criteria; 3. concerns regarding methods used to collect data and appraise studies; 4. Concerns regarding the synthesis and findings). The judgment about risk of bias will be ‘‘high,’’ ‘‘low,’’ or ‘‘unclear’’. The risk of bias assessment from each RCT will be performed by two reviewers, independently; when necessary, a third researcher will solve disagreements. The results will be graphically summarized through graphs.

### Data analysis

The primary analysis will address:

- The completeness of the reporting for each study. Total adherence to the PRSIMA checklist will be calculated (in percentage) as the total number of items described and reported out of the total number of applicable items.
- The total adherence for each item between studies, calculated as (for each item; in percentage) the number of times that one item is described and reported out of the total number of studies that have such item applicable.
- The relationship between PRISMA adherence and risk of bias. ROBIS is not an unidimensional tool, therefore we will not perform a quantitative analysis due to the nature and multidimensionality of both instruments (ROBIS and PRISMA Checklist). This will be a descriptive analysis through descriptive statistics and graphical representation.

Secondary analysis

- The impact of time elapsed (per year) since publication of the articles and total adherence will be evaluated. A regression analysis will be performed to assess the relationship between the adherence to the PRISMA checklist and the year of publication, with use of total adherence (as described above) as dependent variable and publication year (2011-2020) as independent variable.
- In the same way, the relationship between the following characteristics (as independent variables) and the total adherence will be evaluated:
  - Journal quartile
  - Publications modalities (e.g. Open Access vs Subscription vs Hybrid)
  - Published study protocol

## Data Availability

All data are in the manuscript

## ETHICS AND DISSEMINATION

Several studies have shown the positive influence of RGs on the completeness of research reporting^22^. The adherence to these guidelines can result in a published article that contains precise information and that can better allow readers to make informed judgments. By increasing the likelihood that critical information is included in the submitted manuscript, the work of editors and external reviewers can also become more efficient^2^.

Since the publication of the multi-journal editorial of Chan and colleagues^5^, no study investigated the adherence to these RGs in physical therapy research. Therefore, this study will add relevant knowledge that may contribute to improve further the value of rehabilitation research.

This study will also align with the mission statement of the EQUATOR (Enhancing the QUAlity and Transparency of health Research) Network^23^, an international collaboration aiming to enhance the reliability of medical research literature by promoting transparent, accurate reporting of research studies.. A manuscript with results will be prepared and submitted for journal publication upon project completion. The findings of the study will be disseminated at a relevant (inter)national conference. The results of this research will be published in a relevant journal in the rehabilitation category, which has peer review and qualifies physical therapy research and practice. All results of this meta-research study will also be announced at (inter)national scientific events in the area of rehabilitation and research method.

## AUTHORS’ CONTRIBUTIONS

TI, AC, and RO conceived and designed the study protocol. TI, SS, SG, SM and DF designed the draft search strategy. TI, SS, SG, SM, DF, RO and AC were involved in conceptualising the study objectives, providing input into the search strategy, study selection criteria and plans for data extraction. All the authors, including TI, SS, SG, SM, DF, RO and AC, approved the final version of the manuscript.

## REFERENCES

1. Simera I and Altman DG. Writing a research article that is “fit for purpose”: EQUATOR Network and reporting guidelines. Evid Based Med 2009; 14: 132-134. 2009/10/02. DOI: 10.1136/ebm.14.5.132.

2. Golub RM and Fontanarosa PB. Researchers, readers, and reporting guidelines: writing between the lines. Jama 2015; 313: 1625-1626. 2015/04/29. DOI: 10.1001/jama.2015.3837.

3. Altman DG MD. Importance of transparent reporting of health research. In: Altman Dg Mdskfsiwe (ed) Guidelines for Reporting Health Research: A User’s Manual. Wiley, 2014, pp.3– 13.

4. Chalmers I and Glasziou P. Avoidable waste in the production and reporting of research evidence. Obstet Gynecol 2009; 114: 1341–1345. 2009/11/26. DOI: 10.1097/AOG.0b013e3181c3020d.

5. Chan L, Heinemann AW and Roberts J. Elevating the quality of disability and rehabilitation research: mandatory use of the reporting guidelines. The American journal of occupational therapy: official publication of the American Occupational Therapy Association 2014; 68: 127–129. 2014/03/04. DOI: 10.5014/ajot.2014.682004.

6. Chalmers I, Dickersin K and Chalmers TC. Getting to grips with Archie Cochrane’s agenda. British medical journal 1992; 305: 786–788. DOI: 10.1136/bmj.305.6857.786.

7. Paul M and Leibovici L. Systematic review or meta-analysis? Their place in the evidence hierarchy. Clinical microbiology and infection: the official publication of the European Society of Clinical Microbiology and Infectious Diseases 2014; 20: 97–100. 2013/12/21. DOI: 10.1111/1469-0691.12489.

8. Moher D, Tetzlaff J, Tricco AC, et al. Epidemiology and Reporting Characteristics of Systematic Reviews. PLoS medicine 2007; 4: e78. DOI: 10.1371/journal.pmed.0040078.

9. Moher D, Liberati A, Tetzlaff J, et al. Preferred reporting items for systematic reviews and meta-analyses: the PRISMA statement. Annals of internal medicine 2009; 151: 264-269, w264. 2009/07/23. DOI: 10.7326/0003-4819-151-4-200908180-00135.

10. Nawijn F, Ham WHW, Houwert RM, et al. Quality of reporting of systematic reviews and meta-analyses in emergency medicine based on the PRISMA statement. BMC Emergency Medicine 2019; 19: 19. DOI: 10.1186/s12873-019-0233-6.

11. Horsley T, Galipeau J, Petkovic J, et al. Reporting quality and risk of bias in randomised trials in health professions education. Medical education 2017; 51: 61–71. 2016/12/17. DOI: 10.1111/medu.13130.

12. Moher D, Cook DJ, Eastwood S, et al. Improving the quality of reports of meta-analyses of randomised controlled trials: the QUOROM statement. Quality of Reporting of Meta-analyses. Lancet 1999; 354: 1896–1900. 1999/12/10. DOI: 10.1016/s0140-6736(99)04149-5.

13. Page MJ, McKenzie J, Bossuyt P, et al. The PRISMA 2020 Statement: An Updated Guideline for Reporting Systematic Reviews. MetaArXiv 2020. DOI: 10.31222/osf.io/v7gm2.

14. Whiting P, Savović J, Higgins JPT, et al. ROBIS: A new tool to assess risk of bias in systematic reviews was developed. Journal of clinical epidemiology 2016; 69: 225-234. DOI: https://doi.org/10.1016/j.jclinepi.2015.06.005.

15. Tan WK, Wigley J and Shantikumar S. The reporting quality of systematic reviews and meta-analyses in vascular surgery needs improvement: a systematic review. Int J Surg 2014; 12: 1262– 1265. 2014/12/03. DOI: 10.1016/j.ijsu.2014.10.015.

16. Gagnier JJ and Kellam PJ. Reporting and methodological quality of systematic reviews in the orthopaedic literature. Journal of Bone and Joint Surgery - Series A 2013; 95: e771–e777. Review. DOI: 10.2106/JBJS.L.00597.

17. Innocenti T, Salvioli S, Giagio S, et al. Declaration of use and appropriate use of reporting guidelines in high-impact rehabilitation journals is limited: a meta-research study. Journal of clinical epidemiology 2021; 131: 43–50. DOI: 10.1016/j.jclinepi.2020.11.010.

18. Journal Citation Report, https://jcr.clarivate.com (accessed 28/03/2020).

19. Ioannidis JP. The Mass Production of Redundant, Misleading, and Conflicted Systematic Reviews and Meta-analyses. Milbank Q 2016; 94: 485-514. 2016/09/14. DOI: 10.1111/1468-0009.12210.

20. Krnic Martinic M, Pieper D, Glatt A, et al. Definition of a systematic review used in overviews of systematic reviews, meta-epidemiological studies and textbooks. BMC Medical Research Methodology 2019; 19: 203. DOI: 10.1186/s12874-019-0855-0.

21. Random sequence generator, https://www.random.org/sequences/.

22. Turner L, Shamseer L, Altman DG, et al. Consolidated standards of reporting trials (CONSORT) and the completeness of reporting of randomised controlled trials (RCTs) published in medical journals. The Cochrane database of systematic reviews 2012; 11: MR000030. 2012/11/16. DOI: 10.1002/14651858.MR000030.pub2.

23. Altman DG, Simera I, Hoey J, et al. EQUATOR: reporting guidelines for health research. Lancet 2008; 371: 1149-1150. 2008/04/09. DOI: 10.1016/s0140-6736(08)60505-x.

